# COVID-19 outcomes associated with clinical and demographic characteristics in patients hospitalized with severe and critical disease in Peshawar

**DOI:** 10.1101/2022.03.24.22272884

**Authors:** Muhammad Imran, Azharuddin, Muhammad Yousaf, Sajjad Khan, Abdul Jalil Khan, Zafar Iqbal, RahmanUllah Jan, Shahid Khan, Muhammad Salman Khan

**Author notes:** **Corresponding Author** Dr Zafar Iqbal (MBBS, FCPS), Assistant Professor, Pulmonology, Lady Reading Hospital, Peshawar, 25000, Pakistan., (ZI).

## Abstract

**Background:** As a novel disease, understanding the relationship between the clinical and demographic characteristics of coronavirus disease 2019 (COVID-19) patients and their outcome is critical. We investigated this relationship in hospitalized patients in a tertiary healthcare setting.

**Aims/objectives:** To study COVID-19 severity and outcomes in relation to clinical and demographic characteristics of in admitted patients

**Methodology:** In this cross-sectional study, medical records for 1087 COVID-19 patients were reviewed to extract symptoms, comorbidities, demographic characteristics, and outcomes data. Statistical analyses included the post-stratification chi-square test, independent sample t-test, multivariate logistic regression, and time-to-event analysis.

**Results:** The majority of the study participants were >50 years old (67%) and male (59%) and had the following symptoms: fever (96%), cough (95%), shortness of breath (73%), loss of taste (77%), and loss of smell (77%). Regarding worst outcome, multivariate regression analysis showed that these characteristics were statistically significant: shortness of breath (adjusted odds ratio [aOR] 31.3; 95% CI, 11.87–82.53; p < 0.001), intensive care unit (ICU) admission (aOR 28.3; 95% CI,9.0–89.6; p < 0.001), diabetes mellitus (aOR 5.1; 95% CI;3.2–8.2; p < 0.001), ischemic heart disease (aOR 3.4; 95% CI,1.6–7; p = 0.001), nausea and vomiting (aOR 3.3; 95% CI, 1.7–6.6; p = 0.001), and prolonged hospital stay (aOR 1.04; 95% CI, 1.02–1.08; p = 0.001), while patients with rhinorrhea were significantly protected (aOR 0.3; 95% CI, 0.2–0.5; p < 0.001). A Kaplan–Meier curve showed that the symptoms of shortness of breath, ICU admission, fever, nausea and vomiting, and diarrhea increased the risk of mortality.

**Conclusion:** Increasing age, certain comorbidities and symptoms, and direct admission to the ICU increased the risk of worse outcomes. Further research is needed to determine risk factors that may increase disease severity and devise a proper risk-scoring system to initiate timely management.

## Introduction

Severe acute respiratory syndrome coronavirus 2 (SARS-CoV-2) is the cause of coronavirus disease 2019 (COVID-19), a member of the coronavirus family that was responsible for the 2003 SARS epidemic [1]. The World Health Organization (WHO) declared the outbreak to be a Public Health Emergency of International Concern on January 30, 2020, and declared it to be a global pandemic on March 11, 2020 [2]. The virus has affected almost every country, every nation, and all races worldwide. The WHO has reported 269.2 million confirmed cases of COVID-19 and 5.30 million deaths internationally as of December 10, 2021 [3]. Although 8.42 billion vaccine doses were administered worldwide by December 10, only a little more than 64 million doses have been administered in low-income countries [3]. With the emergence of the Omicron variant, the disease is surging again in various parts of the world, thereby posing a challenge to vaccination measures [3]. At the time of this writing, the total number of COVID-19 infections in Pakistan is 1.28 million, with slightly less than 30,000 deaths [4]. Although the vaccination drive was initially slow in Pakistan, its pace is quickening. As of December 8, 2021, 130 million vaccine doses have been administered throughout Pakistan, whereas only 54 million people have been fully vaccinated, defined as having either completed the single-dose vaccine or both doses of the two-dose vaccine [4].

The clinical presentation of COVID-19 is variable [5]. It ranges from asymptomatic infection or mild upper respiratory symptoms in most cases to severe viral pneumonia with respiratory failure [6]. In most infected individuals, no symptoms or only minimal symptoms appear [7]. Symptomatic patients present with variable symptoms. The most common are fever, cough, generalized body aches, shortness of breath, anosmia, and loss of taste. The uncommon symptoms are headache, nausea, and diarrhea [6]. Fever is the most prevalent reported symptom [8]. Continuous fever, shortness of breath, and presence of respiratory and gastrointestinal symptoms make a patient prone to develop severe disease. Moreover, the likelihood of intensive care unit (ICU) admission, mechanical ventilation (invasive and non-invasive), and death is high in such patients. In a study by Rongchen [9] et al., gastrointestinal symptoms were more strongly linked with worse outcomes. In Nigeria, Akin Abayomi [10] et al. found that cough, high-grade fever, and shortness of breath were the most common presenting symptoms. Symptom severity was the most important predictor of death. Among all other symptoms, shortness of breath had a worse prognostic value in terms of mortality. In contrast, isolated loss of the sense of smell and sense of taste was not linked with the development of critical disease and thus had good prognostic value.

Significant risk factors associated with worse outcomes are chronic illness, older age, and genetic predisposition [11]. In a study by Elizabeth [12] et al., older age was the most significant risk factor for death, followed by hematologic malignancies, immunosuppressive drugs, organ transplantation, uncontrolled diabetes mellitus, black race, and obesity. A study from Pakistan by Ambreen Chaudhry [13] et al. found that the risk factors responsible for worse outcomes, defined as ICU admission, invasive mechanical ventilation (IMV), and death, were older age and the presence of comorbidities.

Several studies have reported different mortality rates for patients admitted to different hospital departments, such as the ICU, high-dependency unit, and general ward. Mortality is highest for patients receiving IMV, followed by those managed with continuous positive airway pressure/non-invasive ventilation, high-flow nasal cannula, and oxygen administered via other devices such as a simple face mask and nasal prongs [14]. The outcomes are good for patients who do not require supplemental oxygen and hospital admission [15].

The objective of the current study is to understand the demographic and clinical characteristics of patients with COVID-19 and the associated severity of illness and outcomes. We sought to perform an in-depth analysis of the various factors related to mortality, morbidity, and recovery from COVID-19. In Pakistan, data are very limited on the clinical and demographic characteristics of patients with COVID-19. Therefore, the results of this study will help us tailor our management and preparedness response to the COVID-19 pandemic.

This article was previously presented as an abstract at the 14th Biennial Chest Conference arranged by the Pakistan Chest Society at Karachi, Pakistan, on 4 December, 2021.

## Material and Methods

This cross-sectional study was conducted from July 15, 2020, to July 3, 2021 in Peshawar, Khyber Pakhtunkhwa, Pakistan. All patients admitted to the study hospital were included in our study. Patients who were referred to another hospital or left without medical advice were excluded from our study. Informed written consent was obtained from all patients at admission to the hospital. The analysis was performed using SPSS, version 25 (IBM, Armonk, NY, USA) [16]

The association of various characteristics with mortality was analyzed. We used the post-stratification chi-square test and independent sample t-test to check the statistical significance of qualitative and quantitative characteristics, respectively. A value of p ≤ 0.05 was considered statistically significant. To assess the adjusted association, we used multivariate logistic regression, with mortality as a dependent variable. These predictor variables were included in the final multivariate regression model, with a value of p ≤ 0.25 on the univariate model as the dependent variable. In the final multivariate model, a value of p ≤ 0.05 was considered significant. Time-to-event analysis was conducted to check for the difference in mortality across various groups.

## Results

This study assessed a total of 1087 patients, of whom 449 (41%) were female and 731 (67%) were older than 50 years (Table 1). Most had a fever (96.4%), cough (95.3%), shortness of breath (72.7%), loss of taste (76.6%), and loss of smell (76.5%). Patients presented relatively less commonly with headache (28.1%), rhinorrhea (32.8%), nausea and vomiting (12.8%), and diarrhea (12.1%). Among comorbid conditions, hypertension (29.7%) and diabetes mellitus (24.4%) were more common compared with ischemic heart disease (9%) and end-stage renal disease (1.3%). The average length of hospital stay was 8.33 days. The overall mortality rate was 24.4% (268 patients) and was highest among those admitted directly to the ICU (49 of 56 patients, 88%) versus those admitted directly to the high-dependency unit (210 of 546 patients, 38%) and the general ward (9 of 485, 1.8%).

**Table 1.** Frequency of various characteristics of study participants (n = 1087).

| Characteristics | No. | % |  |
| --- | --- | --- | --- |
| **Age** |  |  |  |
|  | Below 50 years | 356 | 33% |
|  | Above 50 years | 731 | 67% |
| **Gender** |  |  |  |
|  | Female | 449 | 41% |
|  | Male | 638 | 59% |
| **Fever** |  |  |  |
|  | No | 39 | 4% |
|  | Yes | 1048 | 96% |
| **Cough** |  |  |  |
|  | No | 51 | 5% |
|  | Yes | 1036 | 95% |
| **Headache** |  |  |  |
|  | No | 782 | 72% |
|  | Yes | 305 | 28% |
| **Shortness of Breath** |  |  |  |
|  | No | 297 | 27% |
|  | Yes | 790 | 73% |
| **Rhinorrhoea** |  |  |  |
|  | No | 730 | 67% |
|  | Yes | 357 | 33% |
| **Loss of taste** |  |  |  |
|  | No | 254 | 23% |
|  | Yes | 833 | 77% |
| **Diarrhea** |  |  |  |
|  | No | 955 | 88% |
|  | Yes | 132 | 12% |
| **Nausea and Vomiting** |  |  |  |
|  | No | 948 | 87% |
|  | Yes | 139 | 13% |
| **Loss of Smell** |  |  |  |
|  | No | 255 | 23% |
|  | Yes | 831 | 77% |
| **Diabetes Mellitus** |  |  |  |
|  | No | 822 | 76% |
|  | Yes | 265 | 24% |
| **Hypertension** |  |  |  |
|  | No | 764 | 70% |
|  | Yes | 323 | 30% |
| **Ischemic Heart Disease** |  |  |  |
|  | No | 989 | 91% |
|  | Yes | 98 | 9% |
| **End-Stage Renal Disease** |  |  |  |
|  | No | 1073 | 99% |
|  | Yes | 14 | 1% |
| **Days of Hospitalization** |  |  |  |
|  | mean (SD) | 8.33 (7.94) |  |
| **Severity Level** |  |  |  |
|  | Moderate | 146 | 13% |
|  | Severe | 941 | 87% |
| **Unit of Admission** |  |  |  |
|  | HDU / Isolation ward | 1031 | 95% |
|  | ICU | 56 | 5% |
| **Mortality** |  |  |  |
|  | Recovered | 819 | 63% |
|  | Died | 268 | 24% |

Simple stratification (Table 2) showed that mortality was significantly higher in patients older than 50 years (37% vs. 11%) and in those with shortness of breath (41% vs. 2%), diarrhea (60% vs. 24%), nausea and vomiting (60% vs. 23%), diabetes mellitus (62% vs. 16%), hypertension (53% vs. 18%), ischemic heart disease (74% vs. 24%), high level of disease severity (33% vs. 0%), and ICU admission (92% vs. 25%). Although mortality was also higher in female patients (31% vs. 27%) and in those with fever (29% vs. 16%), and end-stage renal disease (100% vs. 28%), the association was not statistically significant. Some characteristics showed a statistically significant protective effect from mortality, such as rhinorrhea (12% vs. 37%), loss of taste (2% vs. 100%), and loss of smell (2% vs. 100%). Length of stay was also significantly higher in patients who died compared with those who survived (11.43 vs. 7.63 days).

**Table 2.** Association of various characteristics with mortality.

| Characteristics |  | Mortality |  |  |  |  |
| --- | --- | --- | --- | --- | --- | --- |
|  |  | Died |  | Recovered |  | *p-value |
|  |  | No. | % | No. | % |  |
| Age |  |  |  |  |  |  |
|  | Below 50 years | 35 | 11% | 277 | 89% | < 0.001 |
|  | Above 50 years | 233 | 37% | 394 | 63% |  |
| Gender |  |  |  |  |  |  |
|  | Female | 118 | 31% | 266 | 69% | 0.217 |
|  | Male | 150 | 27% | 405 | 73% |  |
| Fever |  |  |  |  |  |  |
|  | No | 6 | 16% | 31 | 84% | 0.090 |
|  | Yes | 262 | 29% | 640 | 71% |  |
| **Cough** |  |  |  |  |  |  |
|  | No | 16 | 32% | 34 | 68% | 0.578 |
|  | Yes | 252 | 28% | 637 | 72% |  |
| **Headache** |  |  |  |  |  |  |
|  | No | 197 | 29% | 480 | 71% | 0.543 |
|  | Yes | 71 | 27% | 191 | 73% |  |
| **Shortness of Breath** |  |  |  |  |  |  |
|  | No | 5 | 2% | 290 | 98% | < 0.001 |
|  | Yes | 263 | 41% | 381 | 59% |  |
| **Rhinorrhea** |  |  |  |  |  |  |
|  | No | 232 | 37% | 394 | 63% | < 0.001 |
|  | Yes | 36 | 12% | 277 | 88% |  |
| **Loss of taste** |  |  |  |  |  |  |
|  | No | 254 | 100% | 0 | 0% | < 0.001 |
|  | Yes | 14 | 2% | 671 | 98% |  |
| **Diarrhea** |  |  |  |  |  |  |
|  | No | 191 | 24% | 620 | 76% | < 0.001 |
|  | Yes | 77 | 60% | 51 | 40% |  |
| **Nausea and Vomiting** |  |  |  |  |  |  |
|  | No | 186 | 23% | 617 | 77% | < 0.001 |
|  | Yes | 82 | 60% | 54 | 40% |  |
| **Loss of Smell** |  |  |  |  |  |  |
|  | No | 255 | 100% | 0 | 0% | < 0.001 |
|  | Yes | 12 | 2% | 671 | 98% |  |
| **Diabetes Mellitus** |  |  |  |  |  |  |
|  | No | 113 | 16% | 577 | 84% | < 0.001 |
|  | Yes | 155 | 62% | 94 | 38% |  |
| **Hypertension** |  |  |  |  |  |  |
|  | No | 116 | 18% | 535 | 82% | < 0.001 |
|  | Yes | 152 | 53% | 136 | 47% |  |
| **Ischemic Heart Disease** |  |  |  |  |  |  |
|  | No | 201 | 24% | 647 | 76% | < 0.001 |
|  | Yes | 67 | 74% | 24 | 26% |  |
| **End-Stage Renal Disease** |  |  |  |  |  |  |
|  | No | 264 | 28% | 671 | 72% | - |
|  | Yes | 4 | 100% | 0 | 0% |  |
| **Days of Hospitalization** |  |  |  |  |  |  |
|  | mean (SD) | 11.43 (10.28) |  | 7.63 (6.64) |  | < 0.001 <sup>a</sup> |
| **Severity Level** |  |  |  |  |  |  |
|  | Moderate | 0 | 0% | 136 | 100% | < 0.001 |
|  | Severe | 268 | 33% | 535 | 67% |  |
| **Unit of Admission at Arrival** |  |  |  |  |  |  |
|  | HDU / isolation ward | 219 | 25% | 667 | 75% | < 0.001 |
|  | ICU | 49 | 92% | 4 | 8% |  |

\* p-value represents results of chi-square test
<sup>a</sup> p-value represents results of independent samples t-test

On multivariate regression (Table 3), several characteristics were statistically significant, as follows: shortness of breath (adjusted odds ratio [aOR] 31.3; 95% CI, 11.87–82.53; p < 0.001), ICU admission (aOR 28.3; 95% CI, 9.0–89.6; p < 0.001), diabetes mellitus (aOR 5.1; 95% CI, 3.2–8.2; p < 0.001), ischemic heart disease (aOR 3.4; 95% CI, 1.6–7; p = 0.001), nausea and vomiting (aOR 3.3; 95% CI, 1.7–6.6; p = 0.001) and prolonged hospital stay (aOR 1.04; 95% CI, 1.02–1.08; p = 0.001). These findings that the odds of shortness of breath was 31.3 times, ICU admission was 28.3 times, diabetes mellitus was 5.1 times, ischemic heart disease was 3.4 times, nausea and vomiting was 3.3 times, and prolonged hospital stay was four times higher in patients who died compared with those who survived. Patients with rhinorrhea were significantly protected on a multivariate model (aOR 0.3; 95% CI, 0.2–0.5; p < 0.001).

**Table 3.**
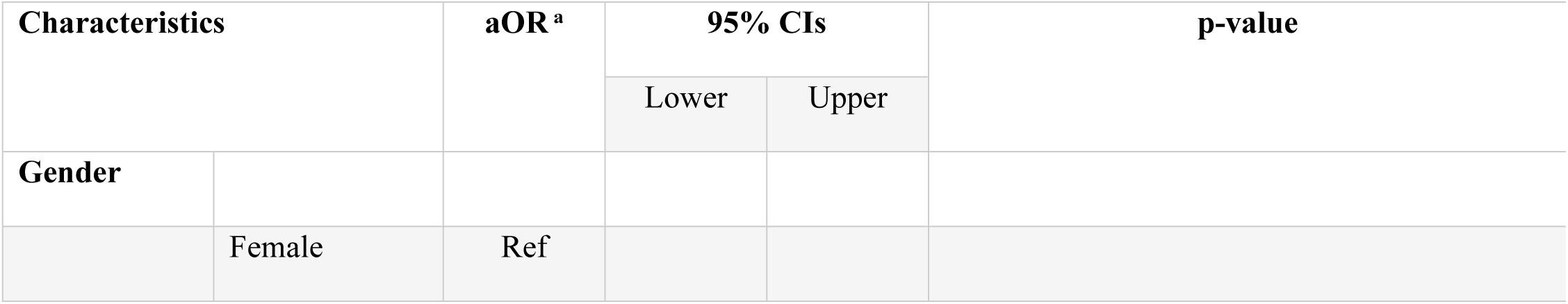
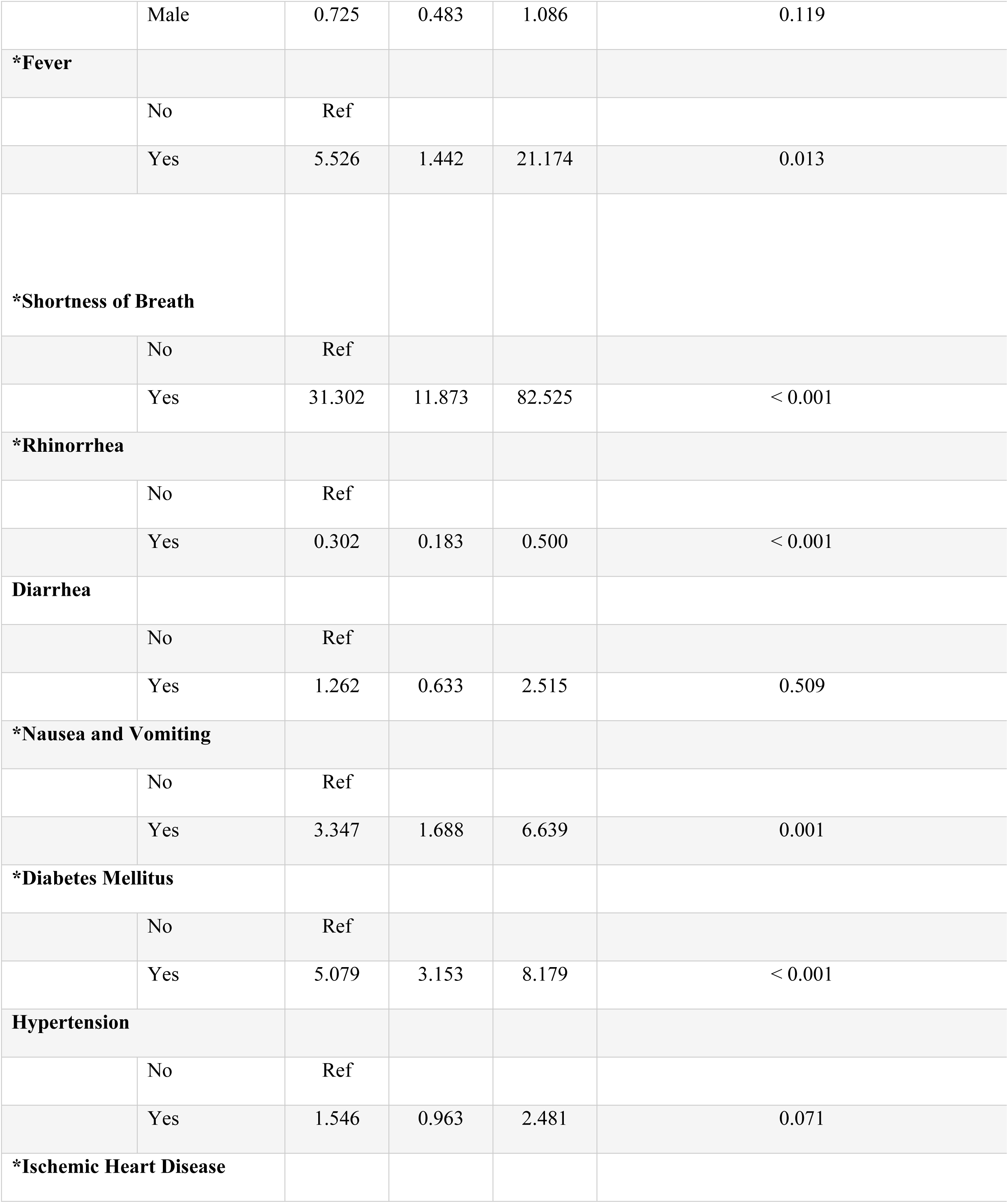
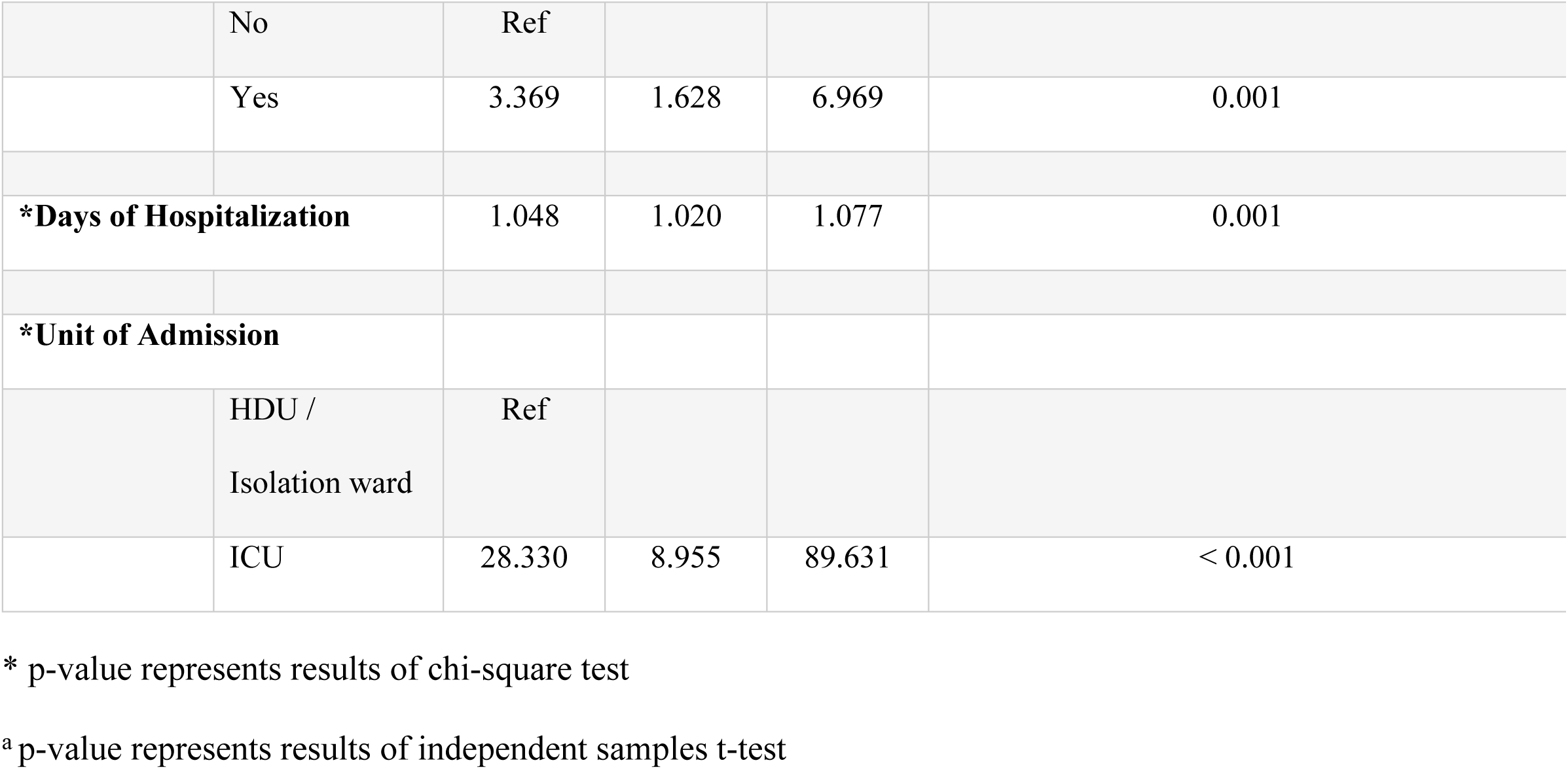
Multivariate logistic regression analysis of various characteristics.

| Characteristics |  | aOR <sup>a</sup> | 95% CIs |  | p-value |
| --- | --- | --- | --- | --- | --- |
|  |  |  | Lower | Upper |  |
| Gender |  |  |  |  |  |
|  | Female | Ref |  |  |  |
|  | Male | 0.725 | 0.483 | 1.086 | 0.119 |
| **\*Fever** |  |  |  |  |  |
|  | No | Ref |  |  |  |
|  | Yes | 5.526 | 1.442 | 21.174 | 0.013 |
| **\*Shortness of Breath** |  |  |  |  |  |
|  | No | Ref |  |  |  |
|  | Yes | 31.302 | 11.873 | 82.525 | < 0.001 |
| **\*Rhinorrhea** |  |  |  |  |  |
|  | No | Ref |  |  |  |
|  | Yes | 0.302 | 0.183 | 0.500 | < 0.001 |
| **Diarrhea** |  |  |  |  |  |
|  | No | Ref |  |  |  |
|  | Yes | 1.262 | 0.633 | 2.515 | 0.509 |
| **\*Nausea and Vomiting** |  |  |  |  |  |
|  | No | Ref |  |  |  |
|  | Yes | 3.347 | 1.688 | 6.639 | 0.001 |
| **\*Diabetes Mellitus** |  |  |  |  |  |
|  | No | Ref |  |  |  |
|  | Yes | 5.079 | 3.153 | 8.179 | < 0.001 |
| **Hypertension** |  |  |  |  |  |
|  | No | Ref |  |  |  |
|  | Yes | 1.546 | 0.963 | 2.481 | 0.071 |
| **\*Ischemic Heart Disease** |  |  |  |  |  |
|  | No | Ref |  |  |  |
|  | Yes | 3.369 | 1.628 | 6.969 | 0.001 |
| **\*Days of Hospitalization** |  | 1.048 | 1.020 | 1.077 | 0.001 |
| **\*Unit of Admission** |  |  |  |  |  |
|  | HDU /<br>Isolation ward | Ref |  |  |  |
|  | ICU | 28.330 | 8.955 | 89.631 | < 0.001 |

\* p-value represents results of chi-square test
<sup>a</sup> p-value represents results of independent samples t-test

The survival analysis comprised 1087 patients. Our analysis observed a cumulated total risk time of 9184 days. The Kaplan–Meier survival plots for the sociodemographic and medical factors that were statistically significant are presented in Figure 1. As can be inferred from the plots, patients with fever (Fig 1A), shortness of breath (Fig 1B), ischemic heart disease (Fig 1C), nausea and vomiting (Fig 1D), diabetes mellitus (Fig 1E), and ICU admission (Fig 1F) were more likely to die compared with patients who did not have these signs and symptoms.

**Fig 1.**
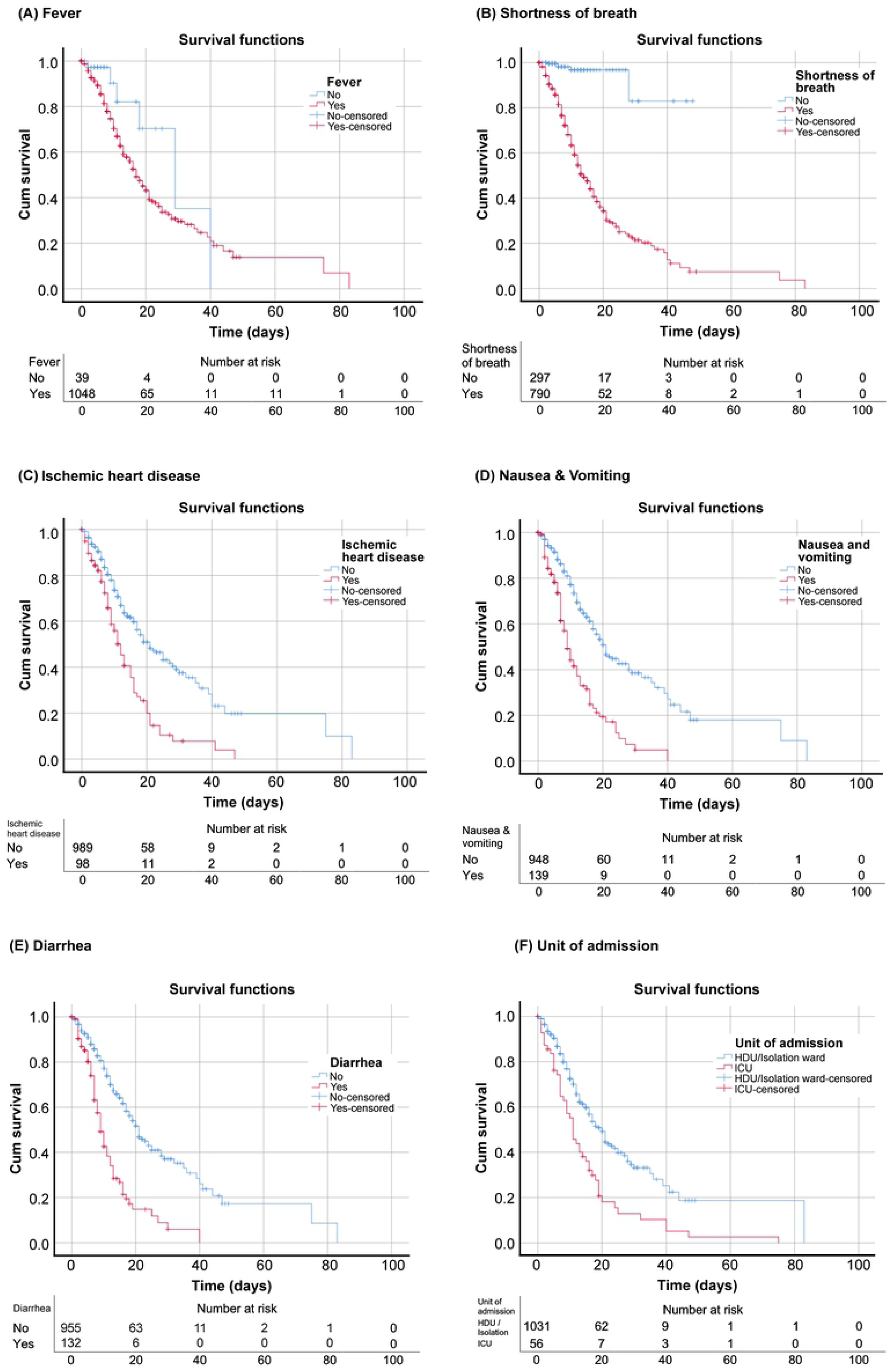
K-M Curves for various characteristics. As shown, patients with fever (A), shortness of breath (B), ischemic heart disease (C), nausea and vomiting (D), diarrhea (E), and ICU admission (F) were more likely to encounter the worst outcome as compared with patients who did not have these characteristics.

## Discussion

The severity of any given disease depends on the population’s demographic and clinical characteristics [17]. Therefore, we aimed to study these factors to understand COVID-19 in detail. This study will help us improve the management steps to prevent severe disease outcomes. Clinicians and scientists are performing many kinds of studies to understand this novel disease, which has killed millions of people. Many studies have already been conducted, but with several drawbacks, such as a small sample size. Although this current investigation is a single-site study, the hospital is an ideal location to examine the COVID-19 cohort because it serves as a referral center for most of the province. This study included 1087 patients with COVID-19 from a single tertiary care institution. Our hospital had an overall mortality rate of 24%, which is comparable with that of most nearby tertiary care hospitals. In a multi-center study conducted by Zia Ul Haq [18] et al. in Peshawar, Pakistan, the inpatient mortality rate in a cohort of 179 patients was 29.1%.

The findings show that certain clinical and demographic factors may predispose patients to worse outcomes. Severe shortness of breath (OR 31) and admission to the ICU on presentation (OR 28), for example, were the strongest predictors of worse outcomes in our analysis, in favor of mortality. Direct admission to the ICU indicates that the patient’s condition is rapidly deteriorating or that the patient arrived at the hospital very late and in critical condition. Furthermore, symptoms such as nausea and vomiting (OR 3.9), fever (OR 3.5), and diarrhea, (OR 1.52) predispose one to worse outcomes as well. In contrast, our study found that rhinorrhea indicated a higher likelihood of recovery, which has not been reported previously. In a survey by Nausheen Nasir [19] et al. in Karachi, Pakistan, disease severity was associated with age older than 60 years (OR 1.92) and having shortness of breath (OR 4.43). In another study, Darbaz A [20] et al. in the United States found that patients presenting with gastrointestinal symptoms had higher hospitalization rates, ICU admission, and intubation. In contrast, Preethi Ramachandran [20] et al. compared the outcomes in two groups of patients with or without gastrointestinal symptoms and discovered that mortality did not differ between cases and controls (41.9% vs. 37.8%; p = 0.68). Secondary outcomes, such as length of stay (7.8 vs. 7.9 days; p = 0.87) and need for mechanical ventilation (29% vs. 26.9%; p = 0.82), also showed no statistically significant differences.

Many studies have found that the presence of comorbidities such as diabetes, hypertension, ischemic heart disease, and end-stage renal disease increases the severity of the disease [22, 23]. In our research, we came to a similar conclusion. According to the survival analysis in our study, patients with diabetes mellitus, hypertension, and ischemic heart disease had a lower chance of survival when admitted to the hospital with COVID-19 than patients without any of these comorbidities. Due to the small sample size, the findings could not be statistically tested because most patients who developed end-stage renal disease in our study were referred to other hospitals with dialysis services. Other authors have reported similar results. Naila Shoaib [24] et al. in Pakistan discovered a link between the severity of COVID-19 and the number and type of comorbidities. As the number of comorbidities increased, so did the disease severity. Patients who were older, had diabetes, or had high blood pressure were more likely to have a poor outcome.

Our study was conducted at a tertiary care hospital with a large sample size for one hospital. The study sample was diverse because the patients were referred from throughout the province to this tertiary care hospital. We evaluated both demographic and clinical characteristics to perform an in-depth analysis.

The limitations of this study are that we did not include patients from other hospitals for comparative analysis. Furthermore, we did not include the response to various treatments, and lastly, the patient’s social characteristics, such as local residence, occupation, and income, were not included in this analysis.

Further research is needed to develop a comprehensive scoring system for COVID-19. The system for scoring should be practical and straightforward to predict the disease severity and the management steps for COVID-19. Both prospective and retrospective analyses from the global north and the global south are needed to identify the critical factors in managing this disease.

## Conclusions

Our findings showed that older age, comorbidities, and direct admission to the ICU increase the risk of worse outcomes. Furthermore, shortness of breath, fever, and gastrointestinal symptoms also increase the risk of worse outcomes. In contrast, patients with isolated upper respiratory symptoms have a lower mortality rate. More research is needed to determine risk factors that increase the risk of disease severity and to devise a proper risk scoring system to initiate timely management.

## Data Availability

Data cannot be shared publicly because of Confidentiality. Data are available from the COVDI-19 Hospital Institutional Data Access(contact via) for researchers who meet the criteria for access to confidential data

## Ethical Approval

Ethical approval for the study was taken from Ethics Committee, Office of Research Innovation & Commercialization (ORIC), Khyber Medical University, Peshawar, bearing number DIR/KMU-EB/DO000667.

Informed written consent was taken from all the participants.

